# Food Insecurity, Cardioprotective Diets, and Mortality Among US Adults

**DOI:** 10.1101/2024.05.07.24307023

**Authors:** Alexander P. Ambrosini, Emily S. Fishman, Sridhar Mangalesh, Golsa Babapour, Zafer Akman, Stephen Y. Wang, Shiwani Mahajan, Karthik Murugiah, Norrisa Haynes, Nihar Desai, Meghana Rao Brito, Jennifer Frampton, Michael G. Nanna

## Abstract

**Importance:** The DASH and Mediterranean diets are cardioprotective. Food insecurity (FI) limits access to healthy food. The relationship between FI, cardioprotective diets, and mortality is unknown.

**Objective:** To assess whether adherence to the DASH or Mediterranean diet affects all-cause and cardiovascular mortality independent of food insecurity. Furthermore, to evaluate if enrollment in the Supplemental Nutrition Assistance Program (SNAP) significantly affects diet quality and interacts with the relationship between cardioprotective diet adherence and mortality.

**Design, Setting, Participants:** A retrospective cohort study using cross-sectional data from the National Health and Nutrition Examination Survey (NHANES) including study cycles 1999-2018 and longitudinal mortality data from the National Death Index (NDI). NHANES is a weighted population-based study that represents the non-institutionalized civilian population in the United States. Adults age ≥18 years with complete food security, dietary recall, and mortality data were included.

**Exposures:** Adherence to DASH or Mediterranean diet and food security status.

**Main outcomes and measures:** All-cause and cardiovascular disease (CVD) related mortality.

**Results:** 51,193 eligible adults were analyzed, representing a weighted population size of 207.1 (200.1 – 214.2) million. Participants with FI were more likely to be younger, female, Black or Hispanic, smokers, obese, lower educational attainment, lower income, and less physically active. FI was a significant predictor of all-cause mortality (p=0.006) but not CVD mortality (p=0.263). After adjustment for multiple covariates and food security status, DASH diet non-adherence significantly predicted all-cause mortality in those with hypertension (p=0.038) and coronary heart disease (CHD) (p=0.001), and CVD mortality in CHD (p=0.041). Mediterranean diet non-adherence significantly predicted all-cause mortality across the entire study population (p=0.005). There was no significant interaction effect between adherence to cardioprotective diets and SNAP enrollment in prediction of all-cause or CVD mortality. Food security was a significant predictor of adherence to the Mediterranean diet (p=0.001). SNAP enrollment was associated with improved adherence to the DASH diet among food insecure individuals (p=0.024), and the Mediterranean diet among food secure individuals (p=0.001).

**Conclusions and relevance:** The relationship between FI and mortality is associated with diet quality. Expanding SNAP eligibility to include food security status may have positive effects on public health.

**Key points:** *Question:* Does adherence to the DASH or Mediterranean diet decrease all-cause and cardiovascular mortality, and is this related to food insecurity status?

*Findings:* Independent of food insecurity, impaired adherence to cardioprotective diets predicted increased risk of all-cause and cardiovascular disease mortality in select patient populations with hypertension or coronary heart disease. SNAP enrollment was not a significant effect modifier in the relationship between mortality and diet adherence, but it did predict increased adherence to the DASH diet in food insecure individuals.

*Meaning:* Our results support the hypothesis that mortality in FI is mediated by impaired diet quality.

## 1. Introduction

Cardioprotective diets including the Dietary Approaches to Stop Hypertension (DASH), DASH-Sodium, and Mediterranean diets have been shown to improve cardiovascular health.^1–7^ However, access to healthy diets is often limited by social determinants of health such as food insecurity (FI).^8^ FI is defined by the United States Department of Agriculture (USDA) as “limited or uncertain access to adequate food,” and in 2022, the estimated prevalence of FI in the United States (US) was 12.8%.^9^ The Supplemental Nutrition Assistance Program (SNAP), previously referred to as the Food Stamp Program, provides food purchasing assistance to low-income households and is the main public health initiative to alleviate FI.^10,11^ FI is associated with an increased risk of cardiovascular disease (CVD), CVD burden, and is an independent predictor of CVD and all-cause mortality in the US.^12–19^ However, the nature of the relationship between FI and mortality is unclear. Some have hypothesized that FI decreases diet quality and thus leads to worse CVD outcomes.^8,20^ Alternate hypotheses include the association of FI with impaired access to care^21–23^ and increased stress and neurohumoral responses implicated in CVD.^24–26^

Understanding the relationship between diet quality, mortality, and FI is important for multiple reasons. Impaired intake of specific food groups has been associated with nearly half of all cardiometabolic deaths,^27^ diet quality is a core tenet of internationally recommended heart-healthy lifestyles,^28–31^ and measures aimed to address FI, such as SNAP, work to improve access to healthy foods.^10^ Changes in adherence to cardioprotective diets, like the DASH and Mediterranean diets, could potentially explain the relationship between FI, and mortality. Specific attention should be paid to those with hypertension (HTN) and coronary heart disease (CHD) as diet has been described as a modifying agent of these diseases.^2,3,5,6,32^ Understanding the role of SNAP enrollment on these factors is key to realizing its potential benefits and shortcomings as a tool to address FI. Prior National Health and Nutrition Examination Survey (NHANES) data demonstrated an association between reduced diet quality and all-cause mortality.^33^

However, FI studies using NHANES data have not found a significant association between diet quality, FI, and CVD mortality,^15^ and fall short of identifying how FI and SNAP affect adherence to cardioprotective diets. Thus, we sought to determine the association of FI and adherence to the DASH and Mediterranean diets with all-cause mortality or CVD mortality. Our secondary objectives were to evaluate if enrollment in SNAP has a significant effect on diet quality and interacts with the relationship between cardioprotective diet adherence and mortality.

## 2. Methods

### 2.1 Study Design and Population

In the present analysis, we used cross-sectional data from NHANES and longitudinal mortality data from participant-level linked mortality files from the National Death Index (NDI). The NHANES is a population-based study by the National Center for Health Statistics (NCHS) on the non-institutionalized civilian population in the United States, assessing the health and nutritional status of adults and children.^34^ The combination of serial cross-sectional data across cycle years has been suggested by the NCHS to improve precision of the nationally representative estimates drawn from NHANES. Participant-level weights provided by NHANES are thus further adjusted by the number of years of data collated for analysis, resulting in average estimates for a calendar year.

Data collection in the NHANES consists of in-home interviews and a physical examination component conducted at mobile examination centers (MEC). Interviews consist of extensive demographic, dietary, health, and socioeconomic-related questions, and the physical exam comprises physiological, medical, dental, and laboratory measurements. For this analysis, data were drawn from twenty years of consecutive NHANES cycles from 1999-2000 through 2017-2018, comprising a total of 101,316 participants. Survey response rates for these years ranged from 49-80% and are adjusted for in the sampling weights provided by NHANES. Inclusion criteria for the analysis were adult participants with complete food security, dietary intake, and NDI linked mortality data. Participants in the pediatric age group (n=36,859) or with insufficient identifying information (n=85) to create a submission record were ineligible for mortality linkage through the NDI. Therefore, only participants age >18 years were included. A total of (n=1,195) participants had missing food security data, and (n=11,984) had missing dietary intake data or zero dietary weights, and were excluded from our analysis. The final resulting sample size was N=51,193 (Figure-1). Informed consent was obtained from all participants by NHANES and the protocol was approved by the NCHS Ethics Review Board. For this analysis, a waiver of review and approval was obtained from the Yale New Haven Hospital institutional review board owing to the anonymized nature of the dataset.

**Figure 1:**
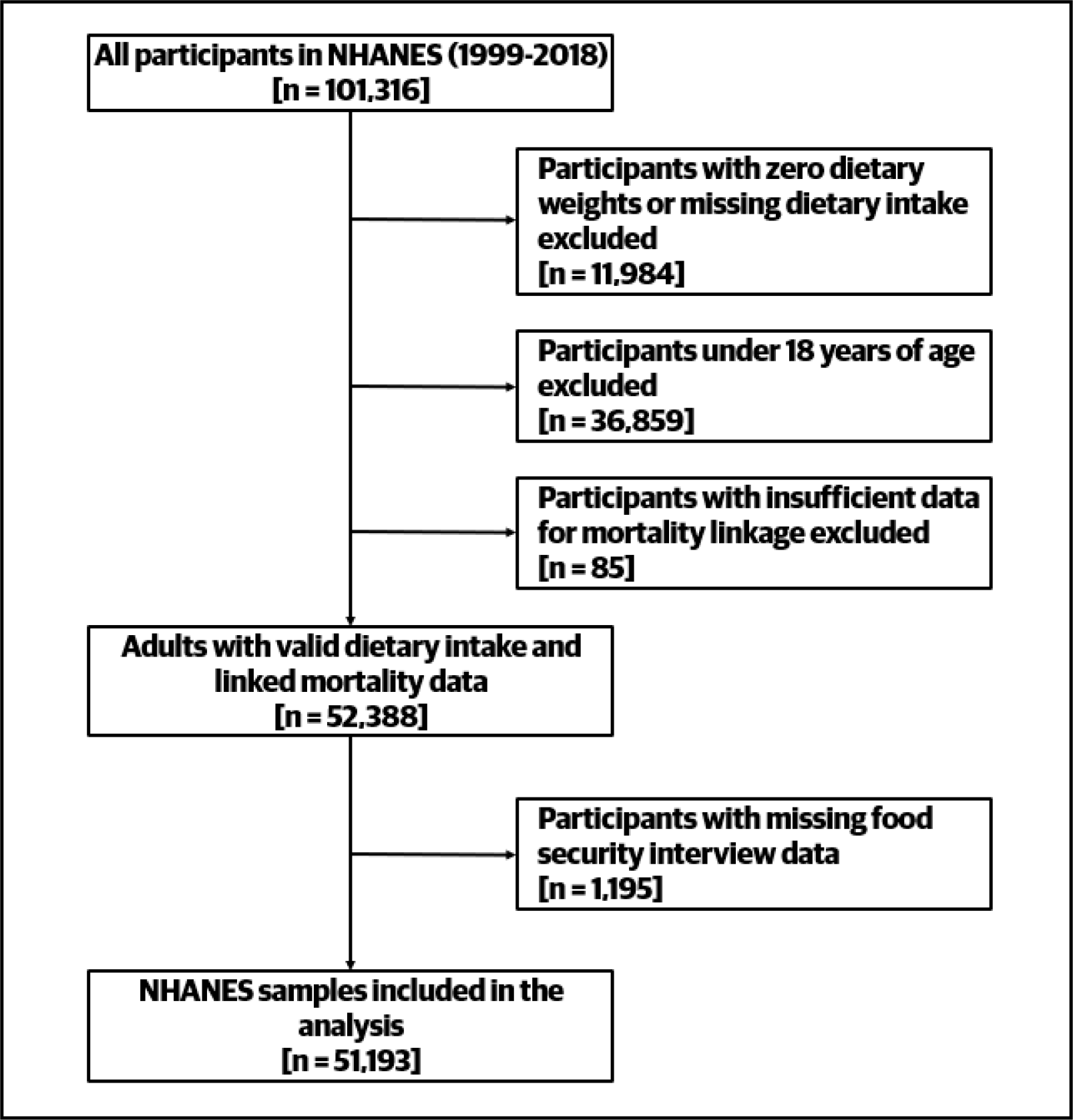
Flowchart depicting study inclusion and exclusion criteria

### 2.2 Mortality Outcomes

Primary outcomes of interest in this analysis were all-cause and CVD mortality. The NCHS provides linked death certificate records through the NDI. The public-use linked mortality files provide mortality follow-up data from the date of survey participation through 31^st^ December 2019. Mortality sources include NDI match, Social Security Administration Information, Center for Medicare and Medicaid Services Information, and death certification match. CVD deaths were those associated with International Classification of Diseases (ICD) codes for acute rheumatic fever, chronic rheumatic heart disease, hypertensive heart disease, hypertensive heart and renal disease, ischemic heart disease, other heat diseases, or cerebrovascular disease.

### 2.3 Food Security Assessment

Food insecurity was defined using participants’ response to the USDA adult food security survey module. In brief, 18 questions regarding food security were asked of households with children under the age of 18, and 10 questions were asked of households with adults only. Food security was defined as 2 or fewer affirmative responses, where 0 affirmatives represented high food security, and 1-2 affirmatives represented marginal food security.^35^ Food insecurity is stratified further as low food security (3-5 affirmatives) and very low food security (6-10 affirmatives).^35^ In this analysis comprising only adult participants, food security was dichotomized based on a cut-point of 2 affirmatives.

### 2.4 Adherence to Cardioprotective Diets

We assessed adherence to the DASH diet and the Mediterranean diet using components of the NHANES dietary intake survey.

#### 2.4.1 DASH Diet

Adherence to the DASH diet was determined using the DASH diet score.^36^ Briefly, the DASH score utilizes dietary goals for nine target nutrients; total fat, saturated fat, protein, fiber, cholesterol, sodium, calcium, potassium, and magnesium. Nutrients are indexed to 1000 kcal of energy intake, apart from macronutrients, which are indexed to total energy intake. Individuals meeting DASH goals for each target are scored one point (for a maximum of 9), and intermediate targets are scored 0.5 points. A cumulative score of 4.5 is used as the cut-point adjudicating adherence to the DASH diet.^36^

#### 2.4.2 Mediterranean Diet

Adherence to the Mediterranean diet was computed using the alternate Mediterranean diet score (aMED), a historically utilized scoring system ranging from 0-9, with higher scores representing greater adherence.^32^ Individuals are assigned points based on their daily intake of certain food groups compared to the median intake of each food group among the entire cohort. Participants with greater-than-median intakes of fruits, vegetables (except potatoes), whole grains, legumes, nuts, fish, and ratio of monounsaturated-to-saturated fat were assigned one point. One point was assigned if alcohol intake was moderate, at 10-25 g/day for men and 5-15 g/day for women, and red or processed meat intake was below the median for the cohort. A total score of 0-3 represents low, 4-5 represents moderate, and 6-9 represents high adherence. For this analysis, participants reporting at least moderate adherence (≥ 4) were considered adherent to the Mediterranean diet.^37,38^

### 2.5 Supplemental Nutrition Assistance Program

Enrollment in SNAP was defined as receiving benefits in the past 12 months. Survey participants reporting individual SNAP use, or SNAP or food stamp use by any member of their household in the previous 12 months were coded as SNAP-enrolled. SNAP enrollment eligibility criteria are established on a state-by-state basis and most commonly require an income-to-poverty ratio less than or equal to 130 percent of the poverty line. Food security status does not affect one’s eligibility for SNAP benefits.

### 2.6 Covariates

Demographic variables included participant age, sex, ethnicity (non-Hispanic white, non-Hispanic black, Mexican American, other Hispanic, other races), ratio of family income-to-poverty, educational attainment (dichotomized into high-school or below and college or above), and marital status (dichotomized into married and not married). Physical activity levels were determined based on self-reported involvement in work and recreational activity, and categorized as vigorous, moderate, or sedentary based on NHANES documentation. Smoking was coded using self-reported history of cigarette smoking, into never, former, and current smoker categories. Body Mass Index (BMI) was calculated using the standard formula. Comorbid conditions were assessed by self-reported diagnosed medical conditions. Diabetes was diagnosed when the participants reported being diagnosed by a health professional and reported taking oral diabetic medications or insulin. Other conditions including CHD, HTN, stroke, and hyperlipidemia were determined by self-reporting diagnoses by a healthcare professional.

### 2.7 Statistical Analysis

In accordance with NHANES analytic guidelines, twenty-year survey weights were calculated and used for all analyses to account for the complex sampling design and resulting unequal selection probabilities and non-response bias.^39^ Stratification and clustering variables provided in the public-use NHANES manual were used in all analyses with MEC-weights or dietary sampling weights as appropriate. Dietary weights additionally account for the day of the week of the 24-hour recall, to adjust for weekday and weekend variation in food intake, in addition to non-response and unequal selection probabilities. Dietary weighting is recommended for use in analyses of dietary intake variables obtained from the 24-hour recall.^39^ We used survey specific descriptive statistics to determine the weighted national estimates for the proportion of participants reporting food insecurity. We examined temporal trends of food security across NHANES cycle years using annual participant-level weights and obtained national estimates for the proportion of food insecurity reported in specific calendar years (Figure-2).

**Figure 2:**
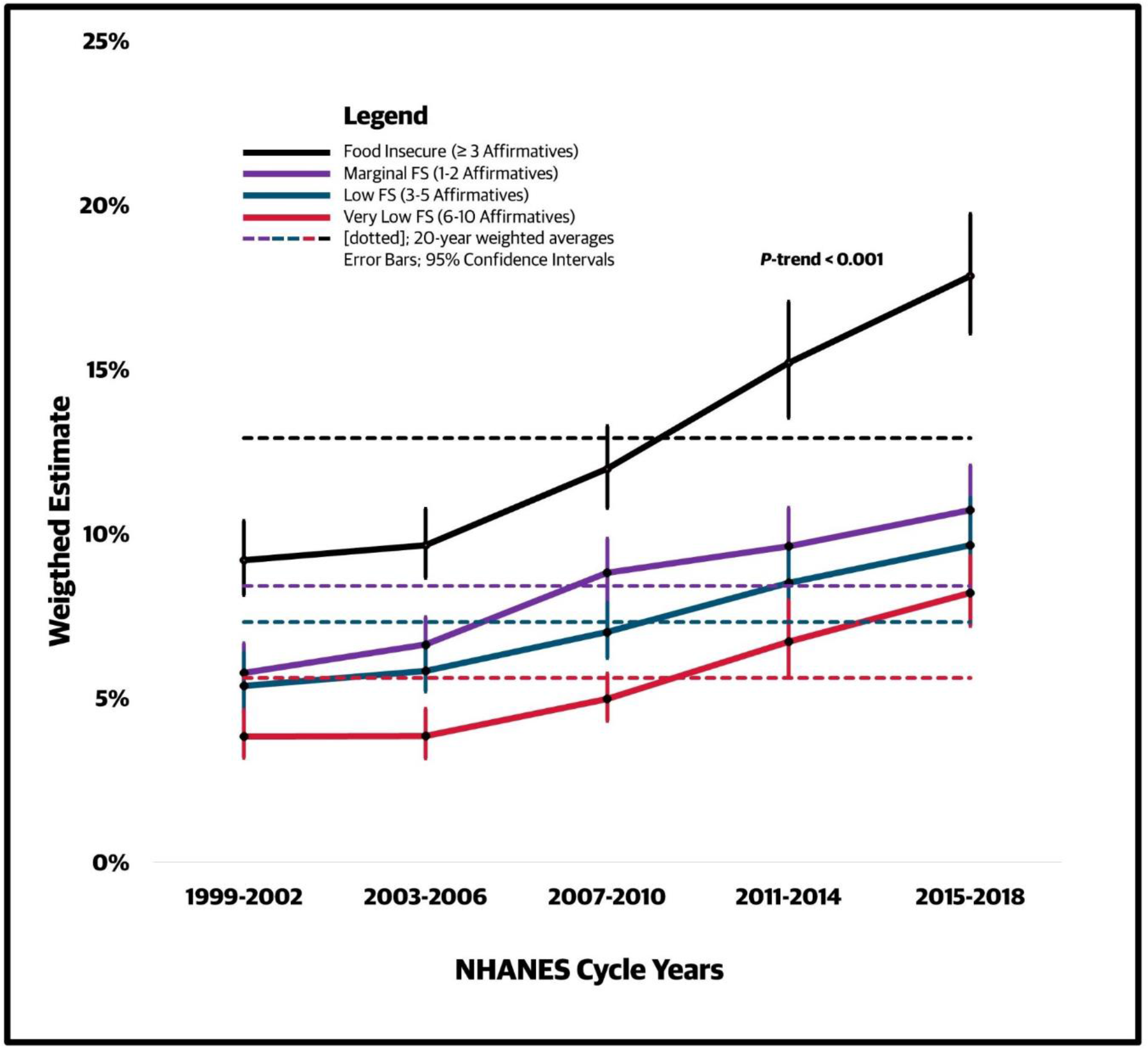
Annual trends in food insecurity

We used a χ^2^ test with the Rao-Scott adjustment to compare categorical variables among participants by food security status. Next, using complex-samples Cox regression models, we examined predictors of all-cause and CVD mortality. Hierarchical models adjusting for demographic, lifestyle and comorbid factors were constructed with food security as a binary categorical variable. Similar multivariable adjusted models were constructed for SNAP enrollment as a predictor of outcomes. We also examined the interaction effect of SNAP enrollment with food security status. Next, multivariable-adjusted models were constructed for DASH and aMED diet scores for predicting both outcome measures in the overall study population and participants that reported pre-existing HTN or CHD. Next, we determined the interaction effect of SNAP enrollment on dietary adherence in multivariable models adjusting for food security status. Lastly, using logistic regression models, we determined the association of food security with adherence to both the cardioprotective diets, and determined if enrollment in SNAP interacts with food security in predicting adherence. The level of significance was set at a *P* = 0.05, and all statistical analyses were performed on SAS v9.4 (SAS Institute, Cary, NC) and SPSS v26.0 (IBM Corp, Armonk, NY).

## 3 Results

### 3.1 Baseline Characteristics

Data from 51,193 eligible adults across the 20-year study period were analyzed, representing a weighted population size of 207.1 (200.1 – 214.2) million. A total of 8,952 (26.8 million; 12.9% [12.3 – 13.6]) participants reported food insecurity. Among them, 5,317 (15.2 million; 7.3% [6.9-7.8]) reported low food security, and 3,635 (11.6 million; 5.6% [5.2-6.0]) reported very low food security. Marginal food security was seen in 5,661 (17.4 million; 8.4% [8.0-8.9]) participants. Temporal trends of food insecurity across NHANES cycles showed a significant increase in the reported frequencies across calendar years (p<0.001) (Figure-2). Food insecurity was noted in 17.3 million (14.5 - 20.1) adults in the study population during 1999-2002 and rose to 39.2 million (34.7 - 43.7) during 2017-2018. Baseline characteristics by food security status are summarized (Supplementary Table-1). Participants reporting food insecurity were more likely to be younger, female, Black or Hispanic, smokers, obese, have lower educational attainment, lower income, and less likely to be physically active.

### 3.2 Food Security and Mortality

A total of 7,561 (weighted; 23.1 million) all-cause mortality events and 1,945 (weighted; 5.7 million) cardiovascular mortality events occurred in the analyzed population. Hazard ratios for food security status in the prediction of all-cause and CVD mortality after adjusting for multiple covariates are presented in (Supplementary Table-2). Diet adherence was not adjusted for in this model. Food insecurity was a significant predictor of all-cause mortality in all three hierarchical models (Model 3 hazard ratio 1.15; 95% CI 1.04 to 1.27). Food insecurity was a significant predictor for cardiovascular mortality when adjusted for age and demographics in Model 1 and 2, but not in Model 3 (hazard ratio 1.11; 95% CI 0.92 to1.34) after adjustment for comorbidities.

### 3.3 SNAP Enrollment and Mortality

There was no significant interaction effect between SNAP enrollment and food security in the prediction of mortality outcomes in the fully adjusted models. Hazard ratios for enrollment in SNAP in the prediction of all-cause and CVD mortality after adjustment for multiple covariates are presented in (Supplementary Table-3). Food security status and diet adherence were not adjusted for in this model. SNAP enrollment remained a significant predictor of all-cause mortality after multivariate adjustment (hazard ratio 1.29; 95% CI 1.13 to 1.48) but not CVD mortality (hazard ratio 1.01; 95% CI 0.81 to 1.25).

### 3.4 Adherence to DASH and Mediterranean Diets and Mortality

Adherence to the DASH diet was noted in 6.2% [5.8 – 6.6] of the study population. The median aMED score was 2.0. Adherence to the Mediterranean diet was noted in 28.5% [27.5 – 29.4] of the study population, and was driven primarily by the consumption of vegetables, fruits, and avoidance of red or processed meats. In the entire population after adjusting for multiple covariates, non-adherence to the DASH diet was not a significant predictor of all-cause (hazard ratio 1.08; 95% CI 0.94 to1.23) or cardiovascular mortality (hazard ratio 1.13; 95% CI 0.86 to 1.48), but non-adherence to the Mediterranean diet was a significant predictor of all-cause mortality (hazard ratio 1.13; 95% CI 1.04 to 1.22) (Supplementary Table-4). Non-adherence to the DASH diet was also a significant predictor of all-cause mortality for those with HTN (hazard ratio 1.20; 95% CI 1.01 to 1.44) and CHD (hazard ratio 1.73; 95% CI 1.25 to 2.40), and for CVD mortality for those with CHD (hazard ratio1.76; 95% CI 1.04 to 3.00). Food security status was not adjusted for in this model.

### 3.5 Adherence to DASH and Mediterranean Diets and Mortality, with Adjustments for Food Security

DASH diet non-adherence was not a significant predictor of all-cause (hazard ratio 1.08; 95% CI 0.94 to 1.23) or CVD mortality (hazard ratio 1.13; 95% CI 0.86 to 1.48) after multivariable adjustment for food security status and other covariates (Table-1). DASH diet non-adherence was associated with all-cause mortality among patients with HTN (hazard ratio 1.21; 95% CI 1.01 to 1.44), and both all-cause (hazard ratio 1.73; 95% CI 1.25 to 2.40) and CVD mortality (hazard ratio 1.76; 95% CI 1.02 to 3.01) among patients with CHD, after adjustment for food security status and other covariates. There was no significant interaction effect of SNAP enrollment on the association between adherence to the DASH diet and mortality outcomes in the fully adjusted models of the entire cohort.

**Table 1:** Adherence to DASH diet and interaction with SNAP enrollment as predictors of all-cause and cardiovascular mortality.

| Model |  | All-Cause Mortality |  | Cardiovascular Mortality |  |
| --- | --- | --- | --- | --- | --- |
|  |  | aHR [95% CI] | P Value | aHR [95% CI] | P Value |
| **DASH diet\*** |  |  |  |  |  |
| DASH Score | Adherent | Ref | - | Ref | - |
|  | Non-Adherent | 1.08 [0.94 – 1.23] | 0.283 | 1.13 [0.86 – 1.48] | 0.378 |
| **Hypertensive** |  |  |  |  |  |
| DASH Score | Adherent | Ref | - | Ref | - |
|  | Non-Adherent | 1.21 [1.01 – 1.44] | 0.038 | 1.20 [0.90 – 1.61] | 0.209 |
| **Coronary Heart Disease** |  |  |  |  |  |
| DASH Score | Adherent | Ref | - | Ref | - |
|  | Non-Adherent | 1.73 [1.25 – 2.40] | 0.001 | 1.76 [1.02 – 3.01] | 0.041 |
| **Interaction with SNAP enrollment** |  |  |  |  |  |
| Total Population | SNAP Enrolled | 1.47 [0.90 – 2.39] | 0.122 | 1.60 [0.53 – 4.79] | 0.403 |
|  | Non-Enrolled | Ref | - | Ref | - |
| DASH Adherent | SNAP Enrolled | 1.74 [0.93 – 3.24] | 0.081 | 1.26 [0.29 – 5.48] | 0.757 |
|  | Non-Enrolled | Ref | - | Ref | - |
| DASH Non-Adherent | SNAP Enrolled | 1.22 [1.04 – 1.43] | 0.012 | 1.01 [0.76 – 1.33] | 0.973 |
|  | Non-Enrolled | Ref | - | Ref | - |
| P Value for interaction |  | 0.438 |  | 0.385 |  |

Complex-samples Cox regression. Aggregated dietary weights were used in accordance with the NHANES weighting module. In brackets: 95% confidence intervals. \*Model adjusted for food security status, age, sex, ethnicity, education, income-poverty ratio, marital status, smoking, physical activity, body mass index, comorbidities, energy intake, & diet adherence. DASH diet adherent; DASH score > 4.5. aHR; adjusted hazard ratio. SNAP; supplemental nutrition assistance program.

Non-adherence to the Mediterranean diet was a significant predictor of all-cause mortality (hazard ratio 1.12; 95% CI 1.04 to 1.22) but not CVD mortality after multivariable adjustment for food security status and other covariates (Table-2). There was no significant interaction effect between adherence to the Mediterranean diet and SNAP enrollment in the prediction of mortality outcomes in the fully adjusted models.

**Table 2:** Adherence to Mediterranean diet and interaction with SNAP enrollment as predictors of all-cause and cardiovascular mortality.

| Model |  | All-Cause Mortality |  | Cardiovascular Mortality |  |
| --- | --- | --- | --- | --- | --- |
|  |  | aHR [95% CI] | P Value | aHR [95% CI] | P Value |
| **Mediterranean Diet\*** |  |  |  |  |  |
| aMED Score | Adherent | Ref | - | Ref | - |
|  | Non-Adherent | 1.12 [1.04 – 1.22] | 0.005 | 1.02 [0.87 – 1.21] | 0.780 |
| **Hypertensive** |  |  |  |  |  |
| aMED Score | Adherent | Ref | - | Ref | - |
|  | Non-Adherent | 1.07 [0.96 – 1.18] | 0.208 | 1.02 [0.84 – 1.24] | 0.802 |
| **Coronary Heart Disease** |  |  |  |  |  |
| aMED Score | Adherent | Ref | - | Ref | - |
|  | Non-Adherent | 1.18 [0.97 – 1.45] | 0.103 | 1.15 [0.82 – 1.60] | 0.416 |
| **Interaction with SNAP enrollment** |  |  |  |  |  |
| Total Population | SNAP Enrolled | 1.33 [1.01 – 1.74] | 0.040 | 1.07 [0.62 – 1.87] | 0.799 |
|  | Non-Enrolled | Ref | - | Ref | - |
| aMED Adherent | SNAP Enrolled | 1.28 [0.93 – 1.76] | 0.122 | 1.01 [0.55 – 1.86] | 0.971 |
|  | Non-Enrolled | Ref | - | Ref | - |
| aMED Non-Adherent | SNAP Enrolled | 1.22 [1.03 – 1.45] | 0.024 | 1.01 [0.74 – 1.38] | 0.935 |
|  | Non-Enrolled | Ref | - | Ref | - |
| P Value for interaction |  | 0.540 |  | 0.788 |  |

Complex samples Cox regression. Aggregated dietary weights were used in accordance with the NHANES weighting module. In brackets: 95% confidence intervals. \*Model adjusted for food security status, age, sex, ethnicity, education, income-poverty ratio, marital status, smoking, physical activity, body mass index, comorbidities, energy intake, & diet adherence. Mediterranean diet adherent; aMED score $\geq 4$ . aMED; alternate Mediterranean diet score. aHR; adjusted hazard ratio. SNAP; supplemental nutrition assistance program.

### 3.6 Food Security and SNAP as a Predictor of Diet Adherence

Food security was a significant predictor of adherence to the Mediterranean diet (hazard ratio 1.30; 95% CI 1.11 to 1.53) but not the DASH diet after multivariable adjustment (Table-3). Similar associations were observed among patients with HTN and CHD. Enrollment in SNAP showed a significant interaction effect with food security status in the prediction of adherence to both diets. SNAP enrollment was associated with improved adherence to the DASH diet among food insecure individuals (hazard ratio 1.73; 95% CI 1.08 to 2.80), and the Mediterranean diet among food secure individuals (hazard ratio 1.66; 95% CI 1.37 to 2.01).

**Table 3:** Food security and SNAP enrollment as predictors of adherence to cardioprotective diets.

| Model |  | DASH Diet Adherence |  | Mediterranean Diet Adherence |  |
| --- | --- | --- | --- | --- | --- |
|  |  | aOR [95% CI] | P Value | aOR [95% CI] | P Value |
| **Food Security Status** |  |  |  |  |  |
| Food Security | Food Secure | 1.17 [0.93 – 1.47] | 0.168 | 1.30 [1.11 – 1.53] | 0.001 |
|  | Food Insecure | Ref | - | Ref | - |
| **Hypertensive** |  |  |  |  |  |
| Food Security | Food Secure | 1.18 [0.81 – 1.71] | 0.380 | 1.49 [1.17 – 1.88] | 0.001 |
|  | Food Insecure | Ref | - | Ref | - |
| **Coronary Heart Disease** |  |  |  |  |  |
| Food Security | Food Secure | 0.68 [0.31 – 1.48] | 0.328 | 2.13 [1.16 – 3.92] | 0.015 |
|  | Food Insecure | Ref | - | Ref | - |
| **Interaction with SNAP Enrollment** |  |  |  |  |  |
| Total Population | SNAP Enrolled | 1.02 [0.81 – 1.27] | 0.252 | 1.69 [1.40 – 2.04] | 0.001 |
|  | Non-Enrolled | Ref | - | Ref | - |
| Food Insecure | SNAP Enrolled | 1.73 [1.08 – 2.80] | 0.024 | 1.08 [0.85 – 1.39] | 0.514 |
|  | Non- Enrolled | Ref | - | Ref | - |
| Food Secure | SNAP Enrolled | 0.84 [0.64 – 1.11] | 0.215 | 1.66 [1.37 – 2.01] | 0.001 |
|  | Non- Enrolled | Ref | - | Ref | - |
| *P Value for interaction* |  | 0.002 |  | 0.005 |  |

Complex-samples logistic regression. Aggregated dietary weights were used in accordance with the NHANES weighting module. In brackets: 95% confidence intervals. Model adjusted for age, energy, ethnicity, income-poverty ratio, and food security; Mediterranean diet adherent - aMED score $\geq 4$ ; DASH diet adherent - DASH score $> 4.5$ . aMED; alternate Mediterranean diet score, aHR; adjusted hazard ratio, DASH; dietary approaches to stop hypertension, SNAP; supplemental nutrition assistance program.

## Discussion

In this retrospective cohort study of a large nationally representative sample of US adults, we found that, independent of food insecurity, impaired adherence to the DASH diet was associated with higher all-cause mortality among participants with HTN and CHD. Non-adherence to the Mediterranean diet was associated with higher all-cause mortality in the entire cohort. While SNAP enrollment did not significantly attenuate these effects, it predicted greater DASH diet adherence in participants with food insecurity. Furthermore, the prevalence of FI significantly increased during the study period, and FI was predictive of decreased adherence to the Mediterranean diet. To our knowledge, this is the first study evaluating the interconnected public health factors of food insecurity, cardioprotective diets, SNAP and their association with mortality outcomes in a single cohort.

The prevalence of FI, socioeconomic, and cardiometabolic profiles of food insecure individuals in this cohort are generalizable to the US population and are similar to those reported in previous FI studies.^9,12,13,15,19,40,41^ Our models demonstrated that FI was associated with increased all-cause mortality. However, this association lost significance in our third model for CVD mortality. This trend is consistent with prior studies using NHANES data which demonstrated increased CVD mortality across adjusted models.^12,15^ These differences are likely attributed to our inclusion of adults age ≥18 years,^15^ differences in covariates, cycle years examined,^12,15^ and use of food security as a reference instead of high food security.^15^ Our selected comorbidities may act as mediators of the association between FI and CVD mortality rather than confounders. It is also possible that the relationship between FI and all-cause and CVD mortality is underappreciated in our analysis because food insecure persons in our cohort were more likely to be young, and therefore less likely to incur all-cause or CVD death.^13–15^

We chose to analyze the DASH and Mediterranean diets because they are commercially ubiquitous, promoted by guidelines, and supported by an abundance of literature demonstrating their positive effects on cardiovascular health.^1–4,28,29,31^ Prior studies have not found an association between FI, diet quality, and mortality.^15^ In contrast, our data support that FI’s negative health outcomes may be related to impaired diet quality. Previous studies have demonstrated an association between decreased diet adherence and worse all-cause and cardiovascular outcomes in specific patient populations.^1–3,5,32^ These associations were notable in our analyses of DASH adherence for those with HTN or CHD, and in Mediterranean adherence for the total population. These relationships were independent of the impact of food security status and other covariates. This novel finding highlights that impaired diet quality is associated with poor health outcomes. In addition, food security was predictive of increased adherence to the Mediterranean diet. This finding, independent of the mortality data presented, is remarkable due to the recognized cardioprotective nature of this diet.

SNAP enrollment was significantly predictive of all-cause mortality. This finding was expected and was previously noted in a similar study using the National Health Interview Survey, an alternate nationally representative sample of US adults.^42^ Prior theories suggest this is due to increased psychosocial burden, medical comorbidities, and impaired diet quality of SNAP participants.^42–44^ Regardless, SNAP has been demonstrated to improve FI,^10^ increase medication adherence,^21^ decrease healthcare expenditures,^45^ and remains the most important food assistance program in the US.^10^ Our results demonstrated that SNAP enrollment was not able to offset the adverse mortality outcomes associated with FI, but was associated with improved DASH diet adherence in participants with FI. Interestingly, SNAP enrollment predicted improved Mediterranean diet adherence for those with food security, but not with FI. This may reflect the restrictive nature of the DASH diet score, which incorporates micro- and macro-nutrient intake. Comparatively, the Mediterranean diet is more inclusive, with points awarded for consumption of diverse food groups. We hypothesize that alleviating the pressure of FI may allow individuals to prioritize diverse food group selection over micro and macro nutrient selection to achieve a balanced diet.

Our study has several implications for public health endeavors. Expanded access to healthy foods should be provided to those with FI and those at risk of experiencing FI. Furthermore, SNAP exists as a measure to address FI but its current eligibility criteria are dependent on income, not food security status. We believe that SNAP eligibility criteria should be expanded to include food insecurity status, as our data demonstrated FI’s negative effects on health are independent of income.^12,15^ Healthy food group incentive programs have been shown as successful ways to leverage SNAP’s financial benefits to promote healthier eating.^46^ This could be an effective way to improve DASH and Mediterranean diet adherence among those enrolled in SNAP. Unfortunately, FI screening is underutilized despite its ability to predict poor health outcomes.^47^ We suggest that all patients, regardless of the clinical setting, be routinely screened for FI using a validated two-question survey.^48^ This has been previously proposed as the “Hunger Vital Sign.”^48–50^ Special attention should be paid to those with HTN and CHD as they were more likely in our study to incur negative health consequences of FI. Over time, DASH adherence has decreased in adults with HTN.^36^ While our study cannot prove this shift is secondary to rising FI, our results suggest that addressing FI and increasing SNAP enrollment may ease this trend. Physicians should continue to promote cardioprotective diets as methods to reduce risk of mortality regardless of a patient’s food security status.^1–3^

Our study has some limitations. NHANES is a cross-sectional study, which limits the ability to comment on causation rather than association between variables. This is especially true as it pertains to the reverse causation between comorbidities and financial burden, which can result in food insecurity.^51,52^ The income-to-poverty ratio in NHANES is top-coded at 500% to protect anonymity. This may have led to loss in data resolution. However, we suspect this loss to be minimal since SNAP eligibility generally starts at or below 130% of the poverty line. Data was gathered in NHANES via survey, which introduces biases related to recall, sampling, and response. Similarly, only one member of each household was surveyed. As food insecurity does not affect each member of the household equally, this may have altered the survey responses. It is also possible that those enrolled in SNAP may under-report food insecurity. Mortality data is limited by the International Classification of Diseases, tenth revision (ICD-10) codes that are classified under cardiovascular mortality. Specifically, CVD-related mortality data includes deaths secondary to diagnoses such as infective endocarditis and cardiac trauma, which do not directly relate to cardiovascular disease processes such as CHD with known pathophysiologic associations to metabolic health.^53^

## Conclusion

Independent of food security status, adherence to the DASH and Mediterranean diets decreases risk of all-cause or CVD mortality in select patient populations. This is particularly evident in those with HTN or CHD. Moreover, food security predicts adherence to the Mediterranean diet, while SNAP is an independent predictor of adherence to the DASH diet in food insecure individuals. These findings will help guide future public health interventions aimed at reducing FI.

## Acknowledgements

### Disclosures

Ambrosini AP: None

Fishman ES: Research related to this study was presented as a moderated poster presentation at ACC.24 in Atlanta, GA.

Mangalesh S: None

Babapour G: None

Akman Z: None

Wang SY: None

Mahajan S: None

Murugiah K: received support from the National Heart, Lung, and Blood Institute of the National Institutes of Health (under award K08HL157727).

Haynes N: None

Desai N: works under contract with the Centers for Medicare and Medicaid Services to develop and maintain performance measures used for public reporting and pay for performance programs. He reports research grants and consulting for Amgen, Astra Zeneca, Bayer, Boehringer Ingelheim, Bristol Myers Squibb, CSL Behring, Cytokinetics, Merck, Novartis, SCPharmaceuticals, and Vifor.

Brito RA: None

Frampton J: reports current research support from the Patient-Centered Outcomes Research Institute (PCORI).

Nanna MG: is a consultant for Merck and HeartFlow, Inc. He reports current research support from the American College of Cardiology Foundation supported by the George F. and Ann Harris Bellows Foundation, the Patient-Centered Outcomes Research Institute (PCORI), the Yale Claude D. Pepper Older Americans Independence Center (P30AG021342), and the National Institute on Aging/National Institutes of Health from R03AG074067.

## Data Availability

All data in this study was generated through the National Health Examination Survey and National Death Index which are publicly available from the National Center for Health Statistics.

https://www.cdc.gov/nchs/nhanes/index.htm

## Notes

### Competing Interest Statement

The authors have declared no competing interest.

### Author Declarations

IRB of the Yale New Haven Hospital

